# A novel esophageal tolerogenic dendritic cell subset

**DOI:** 10.64898/2026.01.18.26344343

**Authors:** Duan Ni, Ralph Nanan

## Abstract

**Background:** Gastroenteric tract requires robust tolerogenic mechanisms to tolerize foreign antigens like foods and microbiota. This is critical to establish the immune homeostasis, which upon disruption, might contribute to a plethora of atopic disorders, including food allergy and eosinophilic esophagitis (EOE). Recently, there was a new subset of tolerizing dendritic cells (tolDCs), PRDM16 tolDC, discovered in the gut of mice and humans, which confers protection against food allergy. Whether an analogous population of it exist in the esophagus is unknown, especially in the context of EOE, another atopic disease associated with dietary antigens.

**Methods:** We thoroughly analyzed the human esophagus cell atlas single cell RNA-seq dataset and the myeloid DC-VERSE dataset, in an attempt to identify and characterize the esophageal counterpart of the intestinal PRDM16 tolDC.

**Results:** We identified the esophageal counterpart of intestinal PRDM16 tolDC as a conventional type II DC subtype expressing *PRDM16*, termed as cDC2C (PRDM16). We demonstrated the similarities between PRDM16 tolDC and cDC2C (PRDM16) regarding their transcriptomic and functional profiles. Importantly, we found that cDC2C (PRDM16) were expanded during EOE and exhibited an anti-inflammatory phenotype, suggesting their protective role in EOE. Notably, these tolerogenic DCs were not found in other atopic diseases beyond the gastroenteric tract.

**Conclusions:** We here defined a novel tolerogenic DC population in human esophagus and demonstrated their implications in the pathophysiology of EOE. These findings would provide novel insights towards the tolerogenic mechanisms along the gastroenteric tract and possess translational relevance for EOE diagnosis and interventions.

---

The gastroenteric tract is continuously exposed to foreign antigens derived from foods and microbiota, requiring robust tolerizing mechanisms to maintain immune homeostasis [1, 2]. Disruption of immune homeostasis is a common trigger to atopic disorders, including food allergy and eosinophilic esophagitis (EOE) [1]. Understanding the basis of gastroenteric tolerance is therefore of substantial translational relevance, offering insights into the pathophysiology of atopic diseases and informing future therapeutic strategies.

Recently, a novel dendritic cell (DC) subset, PRDM16 tolerizing DC (tolDC), was identified in the intestinal tract and shown to play a critical role in maintaining tolerance to gut antigens [2]. These cells promote regulatory T cell (Treg)-mediated tolerance and confer protections against atopic conditions such as food allergy. In human, orthologous PRDM16 tolDC have been identified in gut-draining mesenteric lymph nodes and within the intestinal lamina propria of ileum and colon. Nevertheless, whether this DC population exist in other parts of the gastroenteric tract, specifically the esophagus, remains unknown. Moreover, their potential involvements in eosinophilic esophagitis (EOE), a clinically important atopic disorder commonly triggered by dietary antigens, are yet to be delineated.

Here, we thoroughly analyzed the human esophagus cell atlas [3], which included 37 esophageal biopsies (proximal and distal) from 22 human donors (7 healthy control (Ctrl), 7 remission EOE (Remission), and 8 active EOE (Active)), surveying the transcriptomic profiles of a total of 421312 cells at a single cell level. Overall, the esophagus cell atlas was annotated to 60 cell subsets. Within the atlas, we identified a distinct conventional type II DC subtype, cDC2C, with high expression of *PRDM16* (Figure S1A). This population closely resembled the previously described PRDM16 tolDC [2] (Figure 1A). Comparative analyses of marker genes of esophageal cDC2C (PRDM16) (293 genes) and intestinal PRDM16 tolDC (141 genes) found 35 overlapping genes, including their signature markers like *PRDM16* and *RORC* (Figure 1B). Indeed, within the esophagus atlas, cDC2C (PRDM16) had the highest level of gene set score for PRDM16 tolDC marker genes (Figure 1C). Collectively, these findings indicate that esophegeal cDC2C (PRDM16) likely represent the esophageal counterparts of intestinal PRDM16 tolDC along the digestive tract.

**Figure 1.**
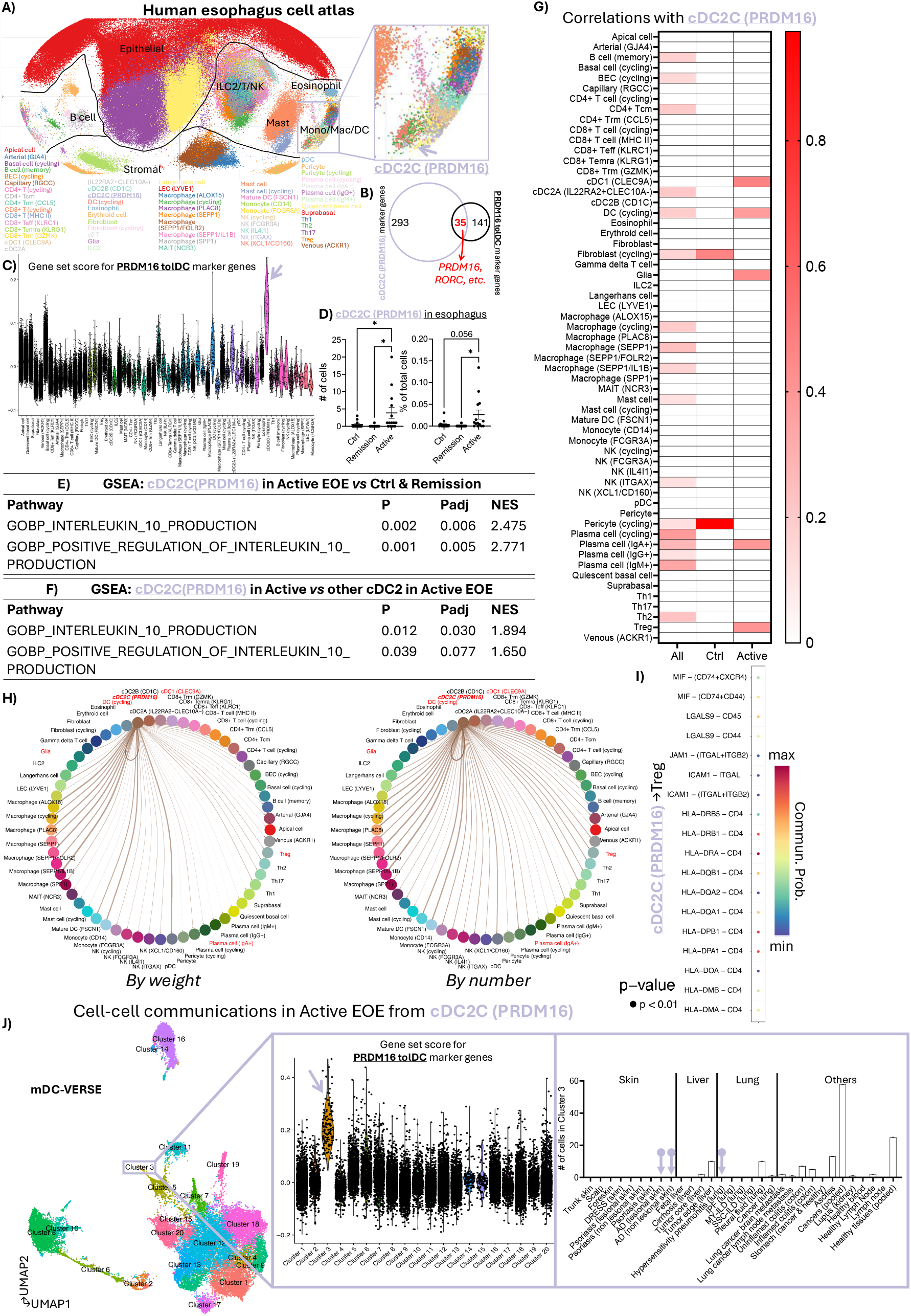
**A.** Overview of the human esophagus cell atlas and cDC2C (PRDM16). **B**. Comparative analysis of the marker genes for esophageal cDC2C (PRDM16) and gastrointestinal PRDM16 tolerizing DC (tolDC). **C**. Gene set score calculations for PRDM16 tolDC marker genes within the human esophagus cell atlas. **D**. Numbers (left) and percentages among all cells (right) of cDC2C (PRDM16) in control esophagus (Ctrl), eosinophilic esophagitis (EOE) remission (Remission) and EOE active (Active) cases. **E**. Gene set enrichment analysis (GSEA) results comparing cDC2C (PRDM16) in Active EOE versus in Ctrl esophagus and Remission EOE. **F**. GSEA results comparing cDC2C (PRDM16) versus other cDC2 subsets in active EOE. **G**. Correlation analyses comparing the percentages of cDC2C (PRDM16) among all cells versus the percentages of other cell populations in all samples (All), control esophagus (Ctrl), and EOE active (Active) cases. The color gradient reflects the R^2^ for correlations and grids in white mean their correlations are not significant. **H**. Cell-cell communication analyses by weight (left) and by number (right) for communication signals originating from cDC2C (PRDM16) to other cell populations during active EOE. **I**. Cell-cell communication analysis results during active EOE for signals originating from cDC2C (PRDM16) and directing to regulatory T cell (Treg). **J**. Overview of the myeloid DC-VERSE (mDC-VERSE) dataset (left), the gene set scores for PRDM16 tolDC marker genes in mDC-VERSE (middle), and the distributions of Cluster 3, resembling cDC2C (PRDM16)/PRDM16 tolDC, across different tissues and conditions, with atopic diseases labelled with arrows. (*: p<0.05; P: p value; Padj: false discovery rate-adjusted p value; NES: normalized enrichment score; BEC: blood vascular endothelial cell; cDC: conventional dendritic cell; ILC: innate lymphoid cell; LEC: lymphatic endothelial cell; MAIT cell: mucosal-associated invariant T cell; NK cell: natural killer cell; pDC: plasmacytoid dendritic cell; Th1 cell: type I T helper cell; Th2 cell: type II T helper cell; DRESS: drug reaction with eosinophilia and systemic symptoms; AD: atopic dermatitis; IPF: idiopathic pulmonary fibrosis; MY-ILD: myositis-associated interstitial lung disease; SSC-ILD: systemic sclerosis-related interstitial lung disease).

In Active EOE, both the absolute numbers and percentages of cDC2C (PRDM16) were significantly increased compared with Ctrl and Remission samples (Figure 1D). No significant differences were found between proximal and distal esophageal regions in active cases (Figure S1B). cDC2C (PRDM16) exhibited a more tolerogenic phenotype in Active EOE than in Ctrl and Remission conditions, as demonstrated by Gene Set Enrichment Analysis (GSEA), showing their upregulation of gene sets associated with anti-inflammatory IL-10 production (Figure 1E). Particularly, in active EOE cases, cDC2C (PRDM16) exhibited a more pronounced anti-inflammatory phenotype than other type II cDC populations, including cDC2A (IL22RA2+CLEC10A-) and cDC2B (CD1C), as evidenced by stronger enrichments of IL-10 production-related gene sets (Figure 1F). Collectively, these findings indicated the anti-inflammatory role of cDC2C (PRDM16) in active EOE, mirroring the tolerizing roles of PRDM16 tolDC in the gut.

Previously, PRDM16 tolDC were found to induce tolerance towards gut antigens through the induction of Treg [2]. Consistent with this, our analysis found that frequencies of cDC2C (PRDM16) were positively correlated with Treg frequencies in Active EOE, which was not found in other states (Figure 1G). To further elucidate their functional interactions, we performed cell-cell communication analyses. In active EOE, cDC2C (PRDM16) exhibited the strongest crosstalk with macrophage populations. Moderate crosstalk was found between cDC2C (PRDM16) and Treg (Figure 1H). Over this process, in addition to HLA-CD4 co-activation, LGALS9 from cDC2C (PRDM16) ligated with CD44 on Treg, enhancing their stability and function [4]. Other immunosuppressive signals were also detected, such as interactions via LFA-1, a heterodimer composed of ITGAL and ITGB2, on Treg [5]; and the migration inhibitory factor (MIF) signals through CD74, CXCR4 and CD44 on Treg [6, 7] (Figure 1I). Collectively, these findings suggest that, in active EOE, cDC2C (PRDM16) may promote anti-inflammatory responses via Treg-mediated mechanisms, similar to their intestinal counterparts.

Given the involvement of cDC2C (PRDM16)/PRDM16 tolDC in two common gastroenteric atopic disorders, namely, food allergy and EOE, we next investigated whether they might also be implicated in other atopic diseases affecting epithelial barriers beyond the gastroenteric tract. To this end, we leveraged the recently published myeloid DC-VERSE (mDC-VERSE) dataset [8], which offers an unprecedented cross-tissue overview of human DC biology. mDC-VERSE integrates 41 published datasets across 13 tissues, covering both healthy and disease states, encompassing a total of 38293 mDCs. Phenograph clustering analysis of mDC-VERSE found 20 distinct clusters, with Cluster 3 emerging as a close proxy of cDC2C (PRDM16)/PRDM16 tolDC (Figure 1J). Notably, among atopic diseases presented in mDC-VERSE that affect epithelial barriers outside the gastroenteric tract, namely, atopic dermatitis (AD) and hypersensitivity pneumonitis, Cluster 3 were absent. This observation suggests that cDC2C (PRDM16)/PRDM16 tolDC, may have a preferential tissue specific role in gastroenteric atopic diseases.

Collectively, to the best of our knowledge, this study provides the first evidence in humans, identifying cDC2C (PRDM16) as the esophageal counterpart of gastrointestinal PRDM16 tolDC. Through in-depth single cell analyses, we demonstrated strong accordance between these populations in both transcriptomic signatures and functional profiles. Importantly, these DC subsets were not found in other atopic conditions affecting epithelial barriers outside the gastroenteric tract, supporting their tissue-specific roles in regulating immune homeostasis.

Clinically, the diagnosis of EOE remains challenging and typically requires invasive endoscopy assessment with esophageal biopsy. If the expansion of cDC2C (PRDM16) observed in esophageal tissue during active EOE is reflected in circulation, this population may present potential diagnostic or disease-monitoring utilities in EOE.

Despite their low abundance in esophageal tissue, cDC2C (PRDM16) exhibit potent anti-inflammatory functional properties, including the induction of Treg and immune tolerance. In this respect, they closely resemble their intestinal counterpart, PRDM16 tolDC, which is known to exert strong Treg-inducing capacity despite limited cellular representation [2]. It would be particular informative to determine the antigen specificity of Treg induced via these tolerizing DCs, especially in light of the well-established association between EOE and specific food allergens.

Intriguingly, these tolerogenic DCs seem to be largely restricted to atopic diseases affecting the gastroenteric tract. This observation warrants further investigations into their potential roles in other gastrointestinal atopic conditions, such as food protein-induced enterocolitis syndrome and eosinophilic gastritis. Moreover, the mechanisms governing their tissue tropism, developmental origins, and differentiation cues remain to be uncovered.

In summary, we identified and characterized a previously unrecognised tolerogenic DC population in human esophagus, cDC2C (PRDM16), which closely mirrors the recently identified gastrointestinal PRDM16 tolDC. Our findings redefine the cellular landscape of EOE by uncovering an intrinsic immune-regulatory axis operating within esophageal tissue, thereby advancing the fundamental understanding of EOE pathophysiology. Beyond mechanistic insights, the delineation of this tolerogenic DC subset establishes a conceptual and translational framework with the potential to inform future biomarker development, disease stratification, and immune-modulatory therapeutic strategies in EOE.

## Data Availability

All data produced in the present study are available upon reasonable request to the authors

## Acknowledgements

This study was supported by the Norman Ernest Bequest Fund and The University of Sydney Faculty of Medicine and Health Bright Ideas Grant 2025.

