## Supplementary Information for "A novel esophageal tolerogenic dendritic cell subset"

### Materials and Methods

#### Data curation

Human esophagus atlas data was obtained from the original publication by Ding et al. (https://singlecell.broadinstitute.org/single_cell/study/SCP1242/eoe-eosinophilic-esophagitis) [1]. mDC-VERSE data was retrieved from the original publication by Mulder et al. (https://github.com/gustaveroussy/FG-Lab) [2].

#### Human esophagus atlas analysis

Human esophagus scRNA-seq data were clustered, annotated and analyzed following the original publication [1] using *Seurat* (v4.3.0) package [3]. Marker genes were identified using the *FindMarkers* function in *Seurat*. Gene set scores were calculated using *AddModuleScore* function in *Seurat* based on the marker genes for PRDM16 tolDC reported in the paper by Fu *et al.* [4]. Comparisons and correlation analysis of different cell populations were carried out with GraphPad PRISM. For correlation analysis, eosinophilic esophagitis (EOE) remission cases were not analyzed, as cDC2C (PRDM16) were only found in one sample. Gene Set Enrichment Analysis (GSEA) and cell-cell communication analysis were run with *fgsea* (v1.20.0) package [5] and *CellChat* (v1.6.1) package [6] respectively, following their corresponding tutorials from the developers.

#### mDC-VERSE analysis

mDC-VERSE data were annotated and analyzed following the original publication [2] using *Seurat* (v4.3.0) package. Gene set scores were calculated using *AddModuleScore* function in *Seurat* based on the marker genes for PRDM16 tolDC reported in the paper by Fu et al. [4]. Comparisons of cell clusters in different conditions were carried out with GraphPad PRISM.

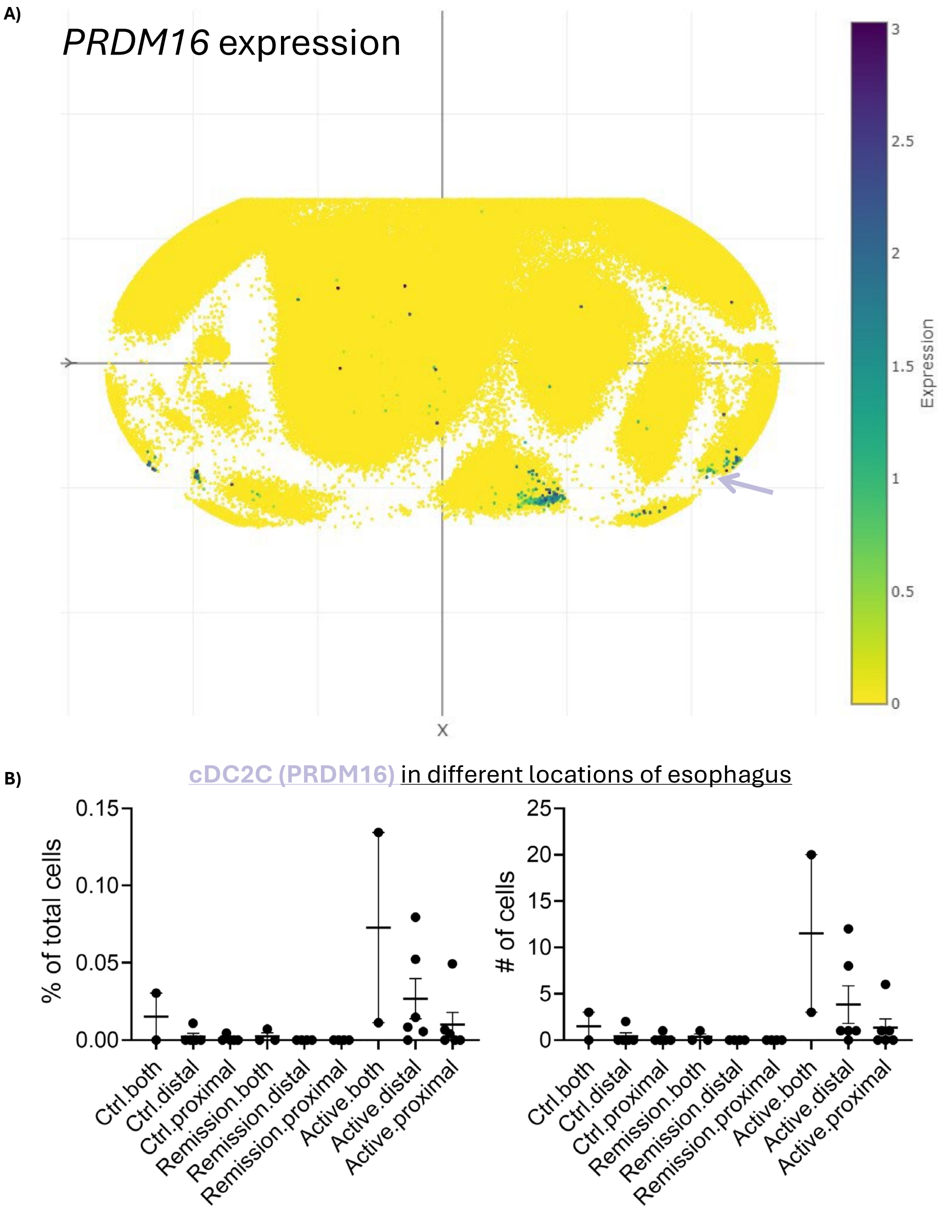


**Supplementary Figure S1. A.** Expression level of *PRDM16* across the human esophagus cell atlas. **B.** Percentages of total cells (left) and numbers (right) of cDC2C (PRDM16) in control esophagus (Ctrl), eosinophilic esophagitis (EOE) remission (Remission) and EOE active (Active) cases in distal esophagus, proximal esophagus, and combined (both, distal and proximal) tissue samples.
